# Psychotherapy remains a human endeavor: Human therapy is valued more than AI-integrated therapy

**DOI:** 10.64898/2026.09.29.26364234

**Authors:** Colin Xu, Thomas T Kim

## Abstract

While Artificial intelligence (AI) is increasingly being integrated into psychotherapy, little is known about how prospective patients evaluate different levels of AI involvement. Participants (n = 1,003) with depression evaluated vignettes describing psychotherapy options, from treatment delivered without AI integration (Human therapist) to treatments involving different degrees of AI integration (Assistive, Collaborative, Fully Autonomous AI). Participants consistently evaluated Human therapists more favorably, reporting a greater likelihood of seeking treatment, less hesitancy, and greater treatment acceptability. Economically, compared with a Human therapist, participants were willing to pay 31.6% less for therapists using Assistive or Collaborative AI and 57.2% less for Fully Autonomous AI. 18-19% of participants indicated that they would refuse treatment involving Assistive or Collaborative AI, compared with 31% for Fully Autonomous AI. Preference for Human over AI therapists was particularly pronounced at higher levels of therapist experience (doctoral/masters vs. peer/trainee). Overall, participants valued psychotherapy delivered by Human therapists more highly than AI integrated psychotherapy. These findings suggest that, despite increasing AI integration in mental health care, prospective patients continue to view psychotherapy as a distinctly human endeavor.

## 1. INTRODUCTION

Digital technologies have rapidly expanded across health care, shaping both how researchers study health and behavior and how clinicians deliver treatment. Digital mental health interventions (DMHIs) refer to mental health treatments delivered through digital technologies, such as mobile applications, web-based platforms, text messaging services, videoconferencing, and chatbots ^1^. More recently, artificial intelligence (AI) has become increasingly prominent within digital health care, with applications in diagnosis ^2^, treatment augmentation ^3^, and the prediction of treatment outcomes ^4^.

Advancements in natural language processing (NLP) and generative AI have contributed to growing interest in chatbots as patient-facing tools for mental health support. For example, a 2026 Pew Research Center survey found that one in ten adults reported using AI chatbots for emotional support or advice ^5^. Similarly, Stade, et al. ^6^ conducted a nationally stratified survey of 1,871 U.S. adults recruited from Prolific, and estimated that 14-18 million adults may have used large language models for mental health purposes in 2024. Users were disproportionately younger, male, and experiencing higher rates of depression and anxiety, and cited treatment cost, limited insurance coverage, and provider unavailability as barriers to traditional care.

Besides fully autonomous AI chatbots being used for mental health care, integration of AI into existing medical treatments has the potential to increase the efficiency of clinical workflows. For example, AI-driven transcription systems have shown promise in converting physician-patient dialogues into actionable, standardized medical notes ^7^. In psychotherapy settings, AI could be used to organize clinical information, reduce documentation burden, summarize treatment-relevant material, or support continuity between sessions. Stade et al. ^8^ described how AI could be integrated at different levels of psychotherapy for mental health, ranging from Assistive AI, to Collaborative AI, to Fully Autonomous AI.

Indeed, mental health services have been exploring AI integration: the online psychotherapy provider, BetterHelp, has publicly described plans to integrate AI into psychotherapy services to “organize data, reduce administrative tasks, or generate text” ^9^. In addition, Spring Health, a digital mental health company that provides employee mental health services, recently evaluated AI-enabled “continuous care” features in combination with psychotherapy ^10^. These optional features included guided intake, session summaries, and session takeaways. Guided intake applied an AI-assisted conversational interface to support structured onboarding before the first psychotherapy session, whereas session summaries and takeaways generated post-session summaries, personalized journaling prompts, and skills-based exercises related to themes from therapy sessions. They found that adding AI-enabled continuous care features to psychotherapy was associated with greater treatment engagement and symptom improvement than psychotherapy alone.

Chatbots are computer programs designed to communicate with users through natural language; they have the potential to deliver psychological support and/or psychoeducation through text- or voice-based interactions ^11^. In a systematic review and meta-analysis of AI-based conversational agents for mental health, Li, et al. ^12^ found that chatbot interventions were associated with significant reductions in symptoms of depression and psychological distress. Reductions were larger for interventions that incorporated generative AI, multimodal or voice-based interaction, mobile applications, or instant messaging platforms. Qualitative feedback from users also suggested that their experiences depended on the perceived quality of the human-AI therapeutic relationship, the quality of intervention content, and communication difficulties. Furthermore, Heinz, et al. ^13^ conducted a randomized controlled trial of Therabot, a generative-AI chatbot trained on expertly written therapist-patient dialogues based on third-wave CBT. In a sample of adults with clinical-level symptoms of major depressive disorder (MDD), generalized anxiety disorder (GAD), and clinically high-risk feeding and eating disorder (CHR-FED), participants in the intervention condition showed greater reductions in depression, anxiety, and weight concerns than those in the waitlist control condition.

In some contexts, AI-generated therapeutic dialogue by chatbots might even resemble human-delivered therapy. Kuhail, et al. ^14^ asked mental health professionals to evaluate transcripts from human-human therapy interactions vs. human-AI interactions. Therapists were unable to reliably distinguish between the two types of transcripts. However, in instances when therapists correctly identified human-human transcripts, they cited features such as empathy, questioning and exploration, and a warm, humanistic tone as evidence of human-delivered therapy.

Despite growing interest in AI-supported behavioral health care, relatively little empirical work has examined stakeholders’ views of AI integration into psychotherapy. Békés and Aafjes-van Doorn ^1^ surveyed clinicians, patients, and a community sample recruited through Prolific about their acceptance of three types of DMHIs: AI-chatbots, AI avatar-based interventions, and teletherapy via videoconferencing. They found that teletherapy was viewed more positively across groups than AI-based DMHIs.

Acceptance of AI chatbots and AI avatars was also strongly associated with general attitudes toward AI. In addition, the community sample reported a higher acceptance of AI chatbots and avatars than did clinicians and patients. In related work, Békés and Aafjes-van Doorn ^15^ used cluster analysis to identify psychological profiles associated with acceptance of AI-based interventions. They found that individuals characterized by high attachment avoidance and low epistemic trust, or a reduced tendency to trust information from others such as a therapist, reported relatively high acceptance of AI-based interventions than teletherapy.

While studies have identified individual characteristics associated with greater acceptance of AI chatbots and AI avatar-based interventions, they have not examined whether acceptance of AI-integration in psychotherapy depends on the level of AI involvement. In addition, relatively little is known about how individuals evaluate AI integration under clinically relevant tradeoffs. For example, preferences for AI-integrated psychotherapy might depend on the therapist’s level of expertise, the severity of the patient’s presenting problem, and the extent to which AI is integrated into treatment. Judgments might also differ depending on individual differences and the outcome being assessed, such as overall treatment preference, willingness to pay, willingness to wait, likelihood of seeking treatment, perceived acceptability, and hesitancy.

Therefore, the present study aimed to examine preferences for AI-integrated psychotherapy by using vignettes that compared a therapist who did not integrate AI into treatment vs. a therapist who did. We varied the following features in the vignettes: the therapist’s level of expertise, the level of AI integration, and the severity of the patient’s presenting problem. Participants were asked to provide judgments of the following: therapist preference, willingness to pay, willingness to wait, likelihood of seeking treatment, perceived acceptability, and hesitancy. In addition, we examined which individual differences were associated with preferences for AI-integrated psychotherapy.

## 2. METHODS

### 2.1 Participants and Procedure

Participants (N=1107) were recruited through Prolific, an online research platform for collecting human-subjects data. This is the same platform used by Stade, et al. ^6^ on their survey of the prevalence of real-world use of large language models for mental health. Prolific has also been found to produce higher-quality data than other commonly used recruitment platforms ^16^. Data collection occurred in several batches between April 10 and April 22, 2026. The approximate cost of the Prolific sample was $8150 ($6100 for participant payments and $2050 for administrative fees).

Eligible participants were adults residing in the United States who indicated through Prolific’s prescreening questions that they had depression. Participants were excluded from analyses if they failed attention or data-quality checks, were flagged by bot-detection procedures, or provided invalid responses. After accounting for missing or incomplete responses and failed attention checks, our final sample (n = 1003) was 55.4% female and ranged in age from 18 to 74 (*M* = 37.15, SD = 11.34). Demographics are reported in **Table S13**.

Participants first provided electronic informed consent. The consent form described the purpose of the study and stated that participation was voluntary and confidential and that participants could withdraw at any time without explanation. The study protocol was approved by the University of Idaho Institutional Review Board (Protocol 26-068). The study was preregistered on the Open Science Framework (OSF): https://osf.io/5ybnj.

### 2.2 Vignettes

Participants completed an online survey in which they were asked to imagine that they were a hypothetical patient seeking psychotherapy for depression and to choose between two possible options for psychotherapy. For each comparison, participants were presented with two vignettes side-by-side, each describing a different psychotherapist. For each vignette pair, one vignette always described a psychotherapy option in which the therapist delivered psychotherapy without the use of AI (hereafter referred to as the “Human” therapist), whereas the other vignette described a psychotherapy option in which AI was integrated into treatment (hereafter referred to as the “AI” therapist). Each psychotherapist was assigned a unique name, which was used in subsequent questions assessing participants’ treatment preferences.

**Table 1** presents an example vignette from the moderate symptom-severity, master’s-level therapist, and Collaborative AI condition (see **Figure S1** for the participant prompts).

**Table 1.** Example vignette comparing psychotherapy with vs. without AI integration.

| Pat | Terry |
| --- | --- |
| Pat is a **masters level therapist** with two years of training in psychotherapy. | Terry is a **masters level therapist** with two years of training in psychotherapy. |
| They deliver psychotherapy via telehealth. | They deliver psychotherapy via telehealth. |
| Pat delivers the full course of psychotherapy by themselves, and creates all their responses during therapy independently. | Terry delivers psychotherapy with a collaborative AI, which creates on-screen messages and responses that Terry can review before saying. |
| Pat also manually creates a plan to structure therapy, creates activity plans and homework assignments, collects and scores assessments, and writes session notes themselves. | The collaborative AI also helps Terry by creating a plan to structure therapy, and creates activity plans and homework assignments, administers and provides feedback on assessments, and writes session notes, which Terry reviews and monitors. |

*Note.* This example shows the moderate symptom severity, master’s-level therapist, and collaborative AI condition.
Before viewing the therapist options, participants in this condition read the following: “Over the past two weeks, you feel depressed and less interested in your usual activities most days, with moderate sleep problems, low energy, poor concentration, and feelings of worthlessness. These symptoms make carrying out your work and daily responsibilities moderately difficult. You have contacted a psychotherapy clinic, which has two therapists available.” Participants then viewed the two therapist options side by side and provided judgments of preference, willingness to pay, willingness to wait, likelihood of seeking therapy, hesitancy, and acceptability.

We used the framework proposed by Stade, et al. ^8^ to construct the levels of AI-integration in psychotherapy. Stade et al. described a continuum of AI integration in psychotherapy, ranging from Assistive AI, to Collaborative AI, to Fully Autonomous AI. In Assistive AI, AI is used as a tool to support clinical providers with tasks that can be offloaded to AI assistants, such as collecting information for patient intakes or assessments, drafting session notes, and summarizing patient worksheets. In Collaborative AI, AI systems take a more active role by providing or suggesting options for treatment planning and therapy content. However, the therapist retains responsibility for final clinical decisions. For example, Collaborative AI might generate session content or between-session assignments for the therapist to review, adapt, and determine whether to use. In Fully Autonomous AI, AI operates as the therapist: it conducts the assessment, selects the appropriate intervention, and delivers the course of therapy – all without direct human oversight.

We further manipulated two additional dimensions: therapist expertise and patient symptom severity. Participants were first randomly assigned to one of four patient-severity conditions: mild, moderate, severe, or severe symptoms with suicidal ideation. Participants were then randomly assigned to receive two of four therapist-expertise conditions: peer support provider, trainee therapist/student-in-training, master’s-level therapist, or doctoral-level psychologist. The descriptions for the severity conditions were informed by the Patient Health Questionnaire-9 (PHQ-9; ^17^). Before each vignette pair, participants were instructed to imagine that they were experiencing the depressive symptoms described and would need to decide between the two psychotherapy options. Participants then read a description of the patient’s depressive symptoms before viewing the vignette pair.

Within their assigned patient-severity and therapist-expertise conditions, participants evaluated all three AI-integration conditions: Assistive AI, Collaborative AI, and Fully Autonomous AI. The order of the AI-integration conditions was randomized across participants. In total, participants provided judgments for six vignette pairs, drawn from 48 possible vignette pairs (4 patient-severity conditions x 4 therapist-expertise conditions x 3 AI-integration conditions). See the Supplementary Materials for the vignette materials; note that the supplement presents the 12 vignette pairs representing all combinations of the four levels of therapist expertise and three levels of AI integration.

### 2.3 Primary outcome measures: Responses to vignettes

After reading each vignette pair, participants reported their overall therapist preference and provided judgments for each therapist regarding the amount they would be willing to pay per session, the length of time they would be willing to wait for an appointment, their likelihood of seeking therapy, their hesitancy about receiving therapy, and the perceived acceptability of receiving therapy.

Therapist preference was assessed using a visual analog scale ranging from a 100-point preference for AI therapist to a 100-point preference for Human therapist, with 0 indicating no preference. (Note that therapist names varied across vignette pairs; “Alex” and “Taylor” are used below as illustrative examples.) For example, participants were asked, “Given that you are feeling moderately depressed, which therapist would you prefer to see?” The scale anchors were “+100, I would prefer Alex” and “+100, I would prefer Taylor.” Participants also rated, on 5-point Likert scales, their likelihood of seeking therapy from each therapist (“How likely are you to seek therapy from Alex?”), their hesitancy about seeking therapy from each therapist (“How hesitant would you be to seek therapy from Alex?”), and the perceived acceptability of receiving therapy from each therapist (“How acceptable would you find receiving therapy from Alex?”). In order to assess the perceived monetary value of psychotherapy, we asked participants to report how much they would be willing to pay to see each therapist (“How much would you be willing to pay out of pocket for a session of therapy with Alex, in U.S. Dollars?”). We also assessed participants’ willingness to wait to see each psychotherapist. For example, participants were asked, “Imagine Alex has a waiting list for therapy, but Taylor has availability right now. How long would you wait to see Alex, instead of starting therapy with Taylor now?” Response options were: “I would not wait – I would start therapy with Taylor immediately,” “Up to 1 week,” “2-3 weeks,” “1 month,” “2 months,” “3 months,” “4-5 months,” “6-9 months,” “9-12 months,” “1 year or longer,” and “I would never choose Alex.” The willingness to pay, willingness to wait, likelihood, hesitancy, and acceptability items were administered twice for each vignette pair, once for each therapist. Therapist preference was assessed once per vignette pair. See **Figure S1** for a sample vignette pair, survey questions, and response options.

### 2.4 Individual differences

Participants reported the following demographic characteristics: age, gender, race/ethnicity, highest level of education, income, employment status, and residential setting (rural, suburban, urban).

They reported the following clinical characteristics: depressive symptom severity, assessed using the PHQ-9 ^17^; anxiety symptom severity, assessed using the Generalized Anxiety Disorder-7 (GAD-7; ^18^); and whether they had ever received help or treatment for a mental health concern.

Participants reported their level of experience with AI tools. They reported their use of AI tools over the past year for several purposes (e.g., searching for information, fun or entertainment). These categories were informed by items from a Pew Research Center survey assessing common uses of chatbots by U.S. adults. Participants also completed measures of AI self-efficacy and AI literacy. AI self-efficacy was evaluated using the Artificial Intelligence Self-Efficacy Scale (AISES; ^19^), which measures individuals’ perceived ability to use and interact with AI technologies. The 22 AISES items are rated on a 7-point Likert scale ranging from strongly disagree to strongly agree, with higher scores indicating greater perceived AI self-efficacy. AI literacy was assessed using the Short AI Literacy Test (AILIT-S; ^20^). The AILIT-S evaluates knowledge across several domains (e.g., what AI is, what AI can do, how AI works, how people perceive AI, and how AI should be used). Each item was scored as correct or incorrect, and total scores were computed by summing the number of correct responses.

Participants also reported the following access-to-care variables: whether they currently had health insurance and the approximate driving time to the nearest hospital with an emergency department.

### 2.5 Statistical analyses

#### 2.5.1 Main effects models

Because the six vignette pairs that each participant encountered varied across the three levels of AI integration (Assistive, Collaborative, and Fully Autonomous) and two of the four therapist experience levels (peer support, trainee, master’s-level, and doctoral-level), responses were nested within participants. Therefore, we used hierarchical linear models to examine the effects of AI integration and therapist experience on treatment judgments while accounting for repeated responses within participants. Pairwise comparisons were conducted using the *emmeans* package, with Bonferroni adjustments applied to multiple comparisons across levels of a factor.

##### 2.5.1.1 Preferences between Human and AI therapists

For models examining therapist preference, the continuous preference rating, ranging from −100 (complete preference for the Human therapist) to +100 (complete preference for the AI therapist), was specified as the dependent variable. Degree of AI integration, defined as the level of AI integration in psychotherapy, was entered as a fixed-effect predictor. A random intercept for participant was included to account for repeated preference ratings across the six vignette pairs.

The model was specified as follows:

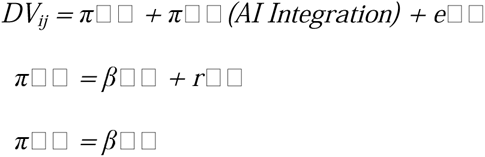

##### 2.5.1.2 Willingness to pay, likelihood of seeking therapy, hesitancy, and acceptability

For these outcomes, participants provided separate ratings for the Human and AI therapists within each vignette pair. Therefore, we included provider type (Human vs. AI therapist) as a dummy-coded fixed-effect predictor, along with degree of AI integration and the interaction between provider type and degree of AI integration. The model was specified as follows:

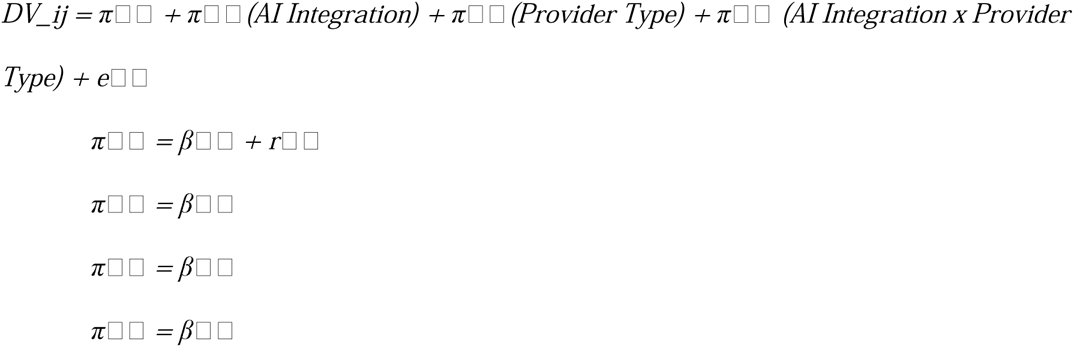

##### 2.5.1.3 Willingness to wait

Because the willingness-to-wait measure included an option to refuse treatment from a provider (“I would never choose [therapist name]”), treatment refusal and wait duration were analyzed separately. We first applied a mixed-effects logistic regression to examine whether participants indicated that they would refuse treatment from a provider. Among responses that did not indicate refusal, we then applied a cumulative link mixed model to examine willingness to wait, treating wait duration as an ordinal outcome.

#### 2.5.2 Moderation analyses

##### 2.5.2.1 Therapist experience

To examine whether therapist experience moderated the effect of AI integration on therapist preference, we added therapist experience (peer support, trainee, master’s level, or doctoral level) and the interaction between therapist experience and degree of AI integration to the models described above.

Within each vignette pair, the Human and AI therapists were presented as having the same level of experience. This allowed us to test whether the effect of AI integration on therapist preference differed according to therapist experience.

##### 2.5.2.2 Vignette depression severity

To examine whether vignette depression severity (the level of depression severity participants were instructed to imagine experiencing when evaluating the psychotherapy options) moderated the effect of AI integration on therapist preference, we added vignette depression severity (mild, moderate, severe, or severe with suicidal ideation) and the interaction between vignette depression severity and degree of AI integration to the models described above.

#### 2.5.3 Individual-difference moderators

To examine whether participant-level characteristics moderated the effect of AI integration on therapist preference, we extended the models described above by including each moderator as a main effect and as an interaction with degree of AI integration. We estimated four grouped models corresponding to demographic characteristics, clinical characteristics, AI experience, and access to care, followed by a combined model that included all moderators simultaneously. Bonferroni adjustments were applied within each model to account for multiple comparisons.

## 3. RESULTS

Our sample comprised 1,003 participants. Participant demographic, clinical, AI experience, and access-to-care characteristics are presented in **Table S13**.

### 3.1 Main effects

#### 3.1.1 Therapist preference

When presented with a choice between the Human and AI therapists, participants strongly preferred the Human therapist versus all AI integrative therapies, including Assistive AI (*b* = −35.83, 95% CI [-39.26, −32.39], *t* = −20.47, *p* < .001), Collaborative AI (*b* = −38.29, 95% CI [-41.72, −34.85], *t* = - 21.88, *p* < .001), and Fully Autonomous AI (*b* = −57.70, 95% CI [-61.13, −54.27], *t* = −32.96, *p* < .001).

Preference for Assistive or Collaborative AI did not significantly differ (*b* = 2.46, 95% CI [-0.54, 5.46], *t* = 1.97, *p* = 0.147), while both Assistive (*b* = 21.87, 95% CI [18.87, 24.88], *t* = 17.45, *p* < .001) and Collaborative (*b* = 19.41, 95% CI [16.41, 22.41], *t* = 15.487, *p* < .001) were significantly preferred compared with Fully Autonomous AI. See **Figure 1**.

**Figure 1.**
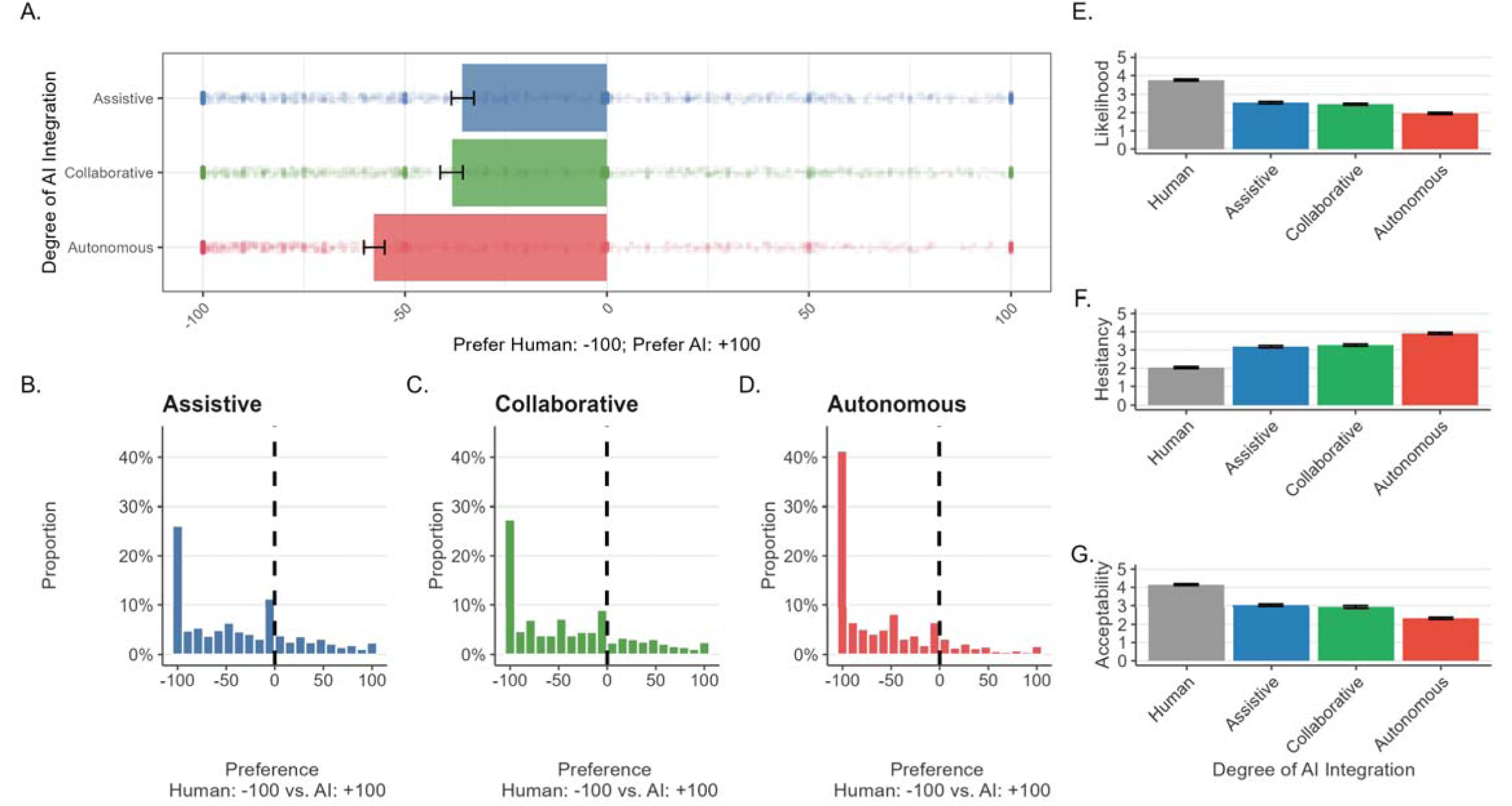
Therapist preference and treatment evaluations by AI integration. **A.** Therapist preference by level of AI integration. “Given that you are feeling [severity] depressed, which therapist would you prefer to see?” Responses ranged from −100 (complete preference for the Human therapist) to +100 (complete preference for the AI therapist), with 0 indicating equal preference. **B-D.** Distributions of therapist preference in the Assistive AI, Collaborative AI, and Fully Autonomous AI conditions, respectively. **E.** Likelihood of seeking therapy from the Human and AI therapists. “How likely are you to seek therapy from [therapist name]?” on a 5-point scale from *Not at all likely* to *Extremely likely*. **F.** Hesitancy about seeking therapy from the Human and AI therapists. “How hesitant would you be to seek therapy from [therapist name]?” on a 5-point scale from *Not at all hesitant* to *Extremely hesitant*. **G.** Acceptability of receiving therapy from the Human and AI therapists. “How acceptable would you find receiving therapy from [therapist name]?” on a 5-point scale from *Totally unacceptable* to *Perfectly acceptable*.

To characterize the distribution of therapist preferences, participants were classified into seven categories based on their mean preference score at each level of AI integration, ranging from Exclusive Human to Exclusive AI (see **Table S14** for category definitions and the complete distribution). Overall, 70.7% of participants preferred the Human therapist in the Assistive AI condition, 73.4% in the Collaborative AI condition, and 82.9% in the Fully Autonomous AI condition.

#### 3.1.2. Willingness to pay

Across the three AI-integration conditions, participants were willing to pay $63.49 (95% CI [$59.51, $67.43], SD = $75.29) for the Human therapist, $43.03 (95% CI [$38.88, $47.19], SD = $70.06) for Assistive, $42.73 (95% CI [$38.58, $46.89], SD = $69.62) for Collaborative, and $27.77 (95% CI [$23.61, $31.92], SD = $63.20) for Fully Autonomous AI. Pairwise comparisons revealed that participants were willing to pay significantly more for the Human therapist than for the AI therapist across all levels of AI integration, with participants willing to pay 31.4% less for Assistive AI (*b* = - $19.70, 95% CI [-$17.52, -$21.88], *t* = −17.71, *p* < .001); 31.8% less for Collaborative AI (*b* = −$19.94, 95% CI [-$17.76, −$22.12], *t* = −17.93, *p* < .001); and 57.2% less for Fully Autonomous AI (*b* = $37.23, 95% CI [-$35.05, −$39.41], *t* = 33.42, *p* < .001). See **Figure 2**. Willingness to pay did significantly not differ between Assistive and Collaborative AI use (*b* = −$0.30, 95% CI [-$2.37, $2.96], *t* = −0.27, *p* = 1.00), while there was a significantly lower willingness to pay for Fully Autonomous AI compared to Assistive (-$15.26, 95% CI [-$12.60, −$17.93], *t* = − 13.71, *p* < .001) and Collaborative (*b* = −$14.96, 95% CI [-$12.30, −$17.63], *t* = −13.44, *p* < .001) AI use.

**Figure 2.**
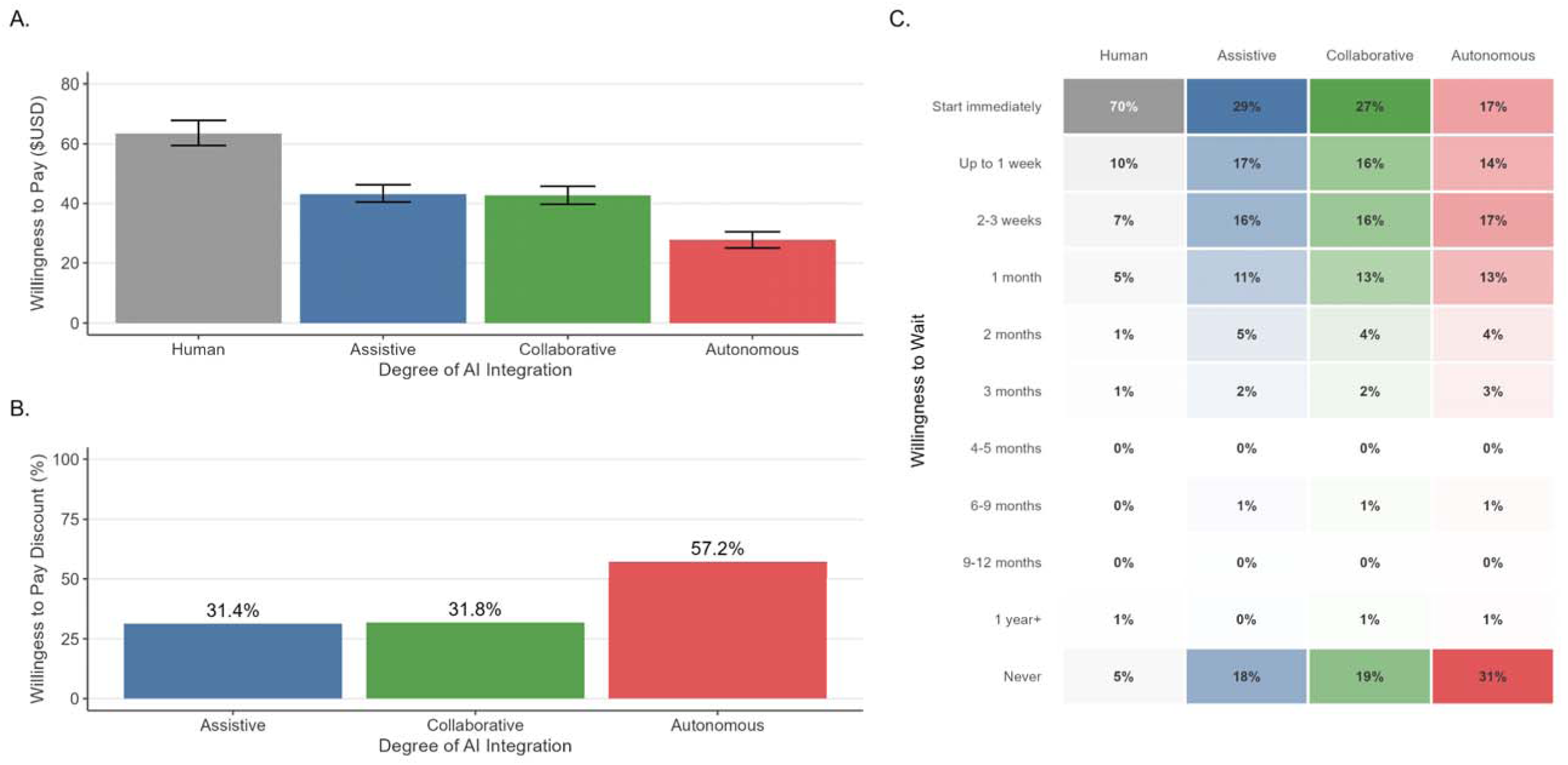
Willingness to pay and willingness to wait by AI integration. **A.** Mean willingness to pay per psychotherapy session for the Human therapist and AI therapists across levels of AI integration. “How much would you be willing to pay out of pocket for a session of therapy with [therapist name] (in U.S. dollars)?” **B.** Percentage reduction in willingness to pay for the AI therapist relative to the Human therapist in the Assistive AI, Collaborative AI, and Fully Autonomous AI conditions. **C.** Distribution of willingness to wait for the Human therapist and AI therapists across levels of AI integration. “If there is a waitlist to see [Therapist A] but no wait to see [Therapist B], how long would you wait to see [Therapist A], instead of starting therapy with [Therapist B] now?” Response options ranged from “I would not wait – I would start therapy with [Therapist B] immediately” to “I would never choose [Therapist B].” The willingness-to-wait question was administered for both the Human and AI therapists.

#### 3.1.3 Willingness to wait

The response options for the willingness-to-wait measure included both wait duration categories (e.g., “Up to 1 week”) and a refusal of treatment option (“I would never choose this therapist”). Because the refusal response could not be meaningfully ordered with the wait duration categories, treatment refusal and wait duration were analyzed separately.

##### Refusal

Overall, 22.4% of participants reported that they would never choose the AI therapist, compared with 5.0% who indicated that they would never choose the Human therapist; refusal of the Human therapist was primarily driven by the peer-support vignettes as described below. Pairwise comparisons revealed that participants were significantly more likely to refuse treatment from the AI therapist than from the Human therapist across all levels of AI integration: Assistive AI (*OR* = 0.106, 95% CI [0.079, 0.142], *z* = −15.04, *p* < .001); Collaborative AI (*OR* = 0.092, 95% CI [0.068, 0.124], *z* = - 15.80, *p* < .001); and Fully Autonomous AI (*OR* = 0.025, 95% CI [0.019, 0.034], *z* = −23.93, *p* < .001).

Refusal rates for the AI therapist did not differ significantly between Assistive (17.8%) and Collaborative (18.5%) AI conditions (*OR* = 1.02, 95% CI [0.84, 1.25], *z* = 0.23, *p* = 1.000), but were significantly higher in the Fully Autonomous AI condition (30.2%) compared to both Assistive (*OR* = 2.12, 95% CI [1.73, 2.59], *z* = 7.28, *p* < .001) and Collaborative (*OR* = 2.07, 95% CI [1.69, 2.53], *z* = 7.02, *p* < .001) conditions. See **Figure 2**.

##### Wait duration

Among participants who indicated that they would be willing to see the therapist, wait duration was analyzed as an ordinal outcome using a cumulative link mixed model. Pairwise comparisons revealed that participants were willing to wait significantly longer for the Human therapist than for the AI therapist across all levels of AI integration: Assistive (*b* = 1.64, 95% CI [1.49, 1.78], *z* = 22.62, *p* < .001), Collaborative (*b* = 1.77, 95% CI [1.63, 1.91], *z* = 24.37, *p* < .001), and Fully Autonomous (*b* = 2.61, 95% CI [2.45, 2.77], *z* = 32.09, *p* < .001). Wait duration did not differ significantly between Assistive and Collaborative AI conditions (*b* = 0.05, 95% CI [−0.06, 0.16], *t* = 0.86, *p* = 1.000), but was significantly higher in the Fully Autonomous condition compared to both Assistive (*b* = 0.60, 95% CI [0.48, 0.71], *t* = 10.22, *p* < .001) and Collaborative (*b* = 0.55, 95% CI [0.43, 0.66], *t* = 9.37, *p* < .001) conditions.

#### 3.1.4. Likelihood, hesitancy, and acceptability

##### Likelihood

Pairwise comparisons revealed that participants reported a significantly greater likelihood of seeking therapy from the Human therapist than from the AI therapist across all levels of AI integration: Assistive AI (*b* = −1.16, 95% CI [-1.23, −1.09], *t* = −31.64, *p* < .001); Collaborative AI (*b* = - 1.28, 95% CI [-1.35, −1.21], *t* = −34.92, *p* < .001); and Fully Autonomous AI (*b* = −1.88, 95% CI [-1.95, - 1.81], *t* = −51.21, *p* < .001). Likelihood of seeking therapy was slightly but significantly lower in the Collaborative compared to the Assistive AI condition (*b* = −0.03, 95% CI [-0.06, −0.01], *t* = −2.90, *p* = .011), and substantially lower in the Fully Autonomous AI condition compared to both Assistive (*b* = - 0.23, 95% CI [-0.26, −0.20], *t* = −13.94, *p* < .001) and Collaborative (*b* = −0.20, 95% CI [-0.23, −0.17], *t* = - 12.58, *p* < .001) conditions. See **Figure 1**.

##### Hesitancy

Pairwise comparisons revealed that participants reported significantly greater hesitancy about seeking therapy from the AI therapist than from the Human therapist across all levels of AI integration: Assistive AI (*b* = 1.13, 95% CI [1.06, 1.20], *t* = 31.49, *p* < .001); Collaborative AI (*b* = 1.23, 95% CI [1.16, 1.30], *t* = 34.16, *p* < .001); and Fully Autonomous AI (*b* = 1.90, 95% CI [1.83, 1.97], *t* = 52.73, *p* < .001). Hesitancy was slightly but significantly greater in the Collaborative compared to the Assistive AI condition (*b* = 0.04, 95% CI [0.01, 0.07], *t* = 2.80, *p* = .015), and substantially greater in the Fully Autonomous AI condition compared to both Assistive (*b* = 0.34, 95% CI [0.30, 0.38], *t* = 16.82, *p* < .001) and Collaborative (*b* = 0.30, 95% CI [0.27, 0.34], *t* = 15.93, *p* < .001) conditions. See **Figure 1**.

##### Acceptability

Pairwise comparisons revealed that participants rated receiving therapy from the Human therapist as significantly more acceptable than receiving therapy from the AI therapist across all levels of AI integration: Assistive AI (*b* = −1.11, 95% CI [-1.18, −1.04], *t* = −32.26, p < .001); Collaborative AI (*b* = −1.21, 95% CI [-1.28, −1.14], *t* = −35.06, *p* < .001); and Fully Autonomous AI (*b* = - 1.85, 95% CI [-1.92, −1.78], *t* = −53.64, *p* < .001). Acceptability was slightly but significantly lower in the Collaborative compared to the Assistive AI condition (*b* = −0.04, 95% CI [-0.06, −0.02], *t* = −3.20, *p* = .004), and substantially lower in the Fully Autonomous AI condition compared to both Assistive (*b* = - 0.32, 95% CI [-0.36, −0.29], *t* = −17.16, *p* < .001) and Collaborative (*b* = −0.28, 95% CI [-0.32, −0.25], *t* = - 15.25, *p* < .001) conditions. See **Figure 1**.

### 3.2 Therapist experience

#### 3.2.1 Therapist preference by therapist experience level

There was a significant main effect of therapist experience on therapist preference, such that a Human therapist was strongly preferred than an AI therapist as therapist experience increased. Specifically, while participants did show a preference for Human over AI even at the peer level (b= - 18.94, 95% CI [-23.57, −14.37] t = −8.04 p <.001), they showed significantly stronger preference for the Human therapist than the AI therapist in the trainee (*b* = −16.21, 95% CI [-21.33, −11.08], *t* = −6.20, *p* < .001), master’s-level (*b* = −24.18, *t* = −9.51 [-29.16, −19.19], *p* < .001), and doctoral-level (*b* = −26.23, 95% CI [-31.38, −21.08], *t* = −9.99, *p* < .001) conditions. See **Figure 3**.

**Figure 3.**
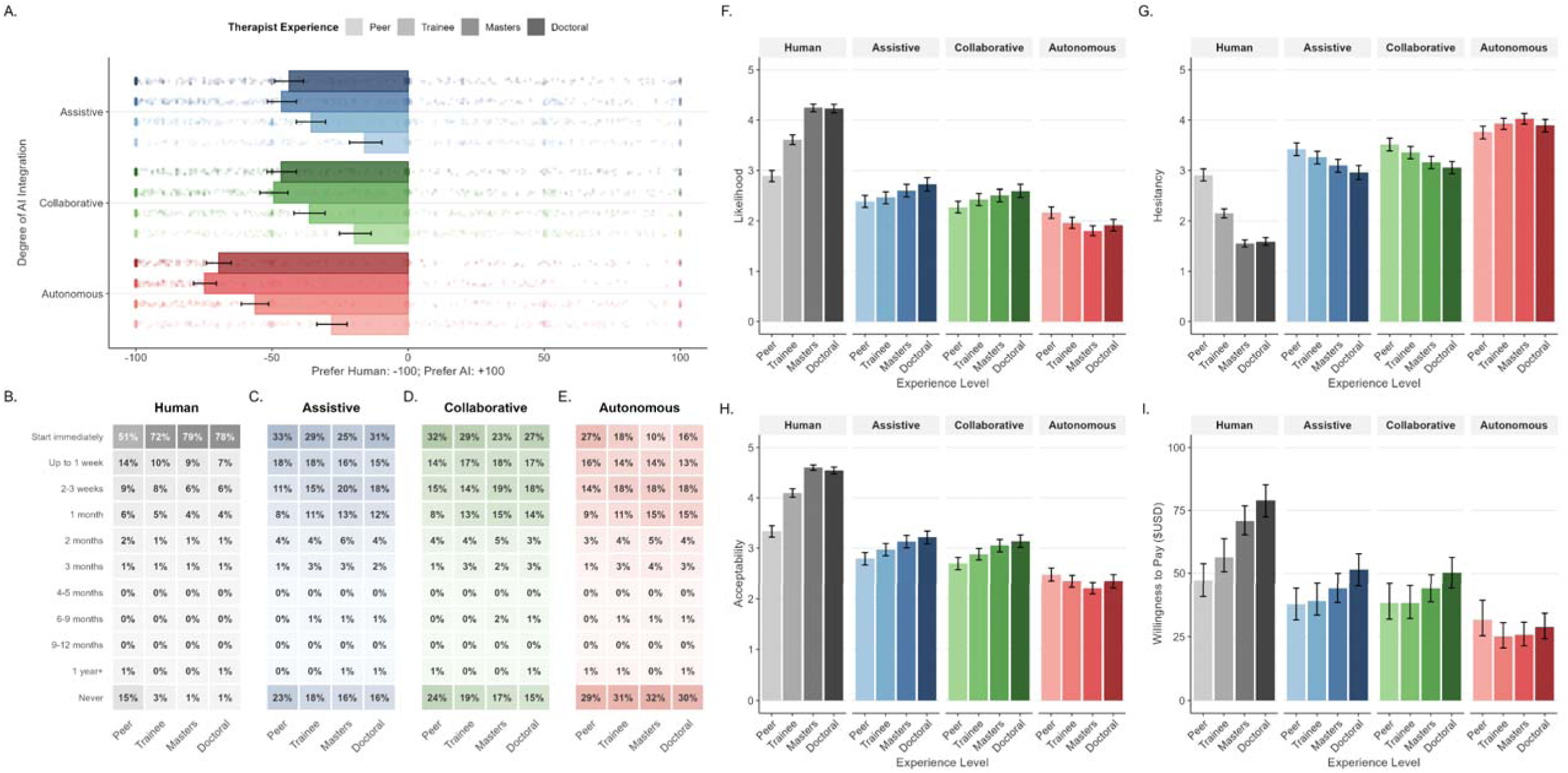
Therapist preference and treatment evaluations by therapist experience and AI integration. **A.** Therapist preference across levels of AI integration and therapist experience. Therapist experience levels were peer support, trainee, master’s level, and doctoral level. Therapist preference ranged from −100 (complete preference for the Human therapist) to +100 (complete preference for the AI therapist), with 0 indicating equal preference. **B-E.** Distributions of willingness-to-wait responses across therapist experience levels for the Human therapist, Assistive AI, Collaborative AI, and Fully Autonomous AI conditions, respectively. **F.** Likelihood of seeking therapy across therapist experience levels. **G.** Hesitancy about seeking therapy across therapist experience levels. **H.** Acceptability of receiving therapy across therapist experience levels. **I.** Mean willingness to pay (US Dollars) per psychotherapy session across therapist experience levels.

Examining AI integration within each therapist-experience level, preference for the Human therapist was significantly stronger in the Fully Autonomous AI condition than in the Assistive AI condition at the peer support (*b* = 11.85, 95% CI [7.03, 16.67], *z* = 4.82, *p* < .001), trainee (*b* = 20.57, 95% CI [15.84, 25.30], *z* = 8.53, *p* < .001), master’s (*b* = 28.29, 95% CI [23.75, 32.83], *z* = 12.22, *p* < .001), and doctoral (*b* = 25.88, 95% CI [21.08, 30.68], *z* = 10.56, *p* < .001) levels. Preference did not significantly differ between the Assistive and Collaborative AI conditions at any level of therapist experience (all *p*s > .05).

#### 3.2.2 Willingness to pay by therapist experience level

Participants were willing to pay significantly more for the Human therapist than for the AI therapist across all 12 combinations of AI integration and therapist experience (all *p*s < .001). At the peer-support level, participants were willing to pay 22% less for the AI therapist in the Assistive AI condition than for the Human therapist ($34.95 vs. $44.88, *b* = $9.93, 95% CI [5.62, 14.24]); 18% less in the Collaborative AI condition ($35.85 vs. $43.66, *b* = $7.81, 95% CI [3.51, 12.11]); and 35% less in the Fully Autonomous AI condition ($29.40 vs. $45.41, *b* = $16.01, 95% CI [11.69, 20.33]; all *p*s < .001).

At the trainee level, participants were willing to pay 29% less for the AI therapist in the Assistive AI condition than for the Human therapist ($40.87 vs. $57.63, *b* = $16.76, 95% CI [12.52, 21.00]); 31% less in the Collaborative AI condition ($39.98 vs. $57.63, *b* = $17.65, 95% CI [13.41, 21.89]); and 55% less in the Fully Autonomous AI condition ($26.77 vs. $59.08, *b* = $32.31, 95% CI [28.07, 36.55]; all *p*s < .001).

At the master’s level, participants were willing to pay 36% less for the AI therapist in the Assistive AI condition ($45.69 vs. $71.25, *b* = $25.56, 95% CI [21.49, 29.63]); 36% less in the Collaborative AI condition ($45.68 vs. $70.98, *b* = $25.30, 95% CI [21.23, 29.37]); and 63% less in the Fully Autonomous AI condition ($27.24 vs. $74.56, *b* = $47.32, 95% CI [43.25, 51.39]; all *p*s < .001).

At the doctoral level, participants were willing to pay 34% less for the AI therapist in the Assistive AI condition than for the Human therapist ($50.34 vs. $76.27, *b* = $25.93, 95% CI [21.63, 30.23]); 37% less in the Collaborative AI condition ($49.13 vs. $77.56, *b* = $28.43, 95% CI [24.13, 32.73]); and 65% less in the Fully Autonomous AI condition ($27.75 vs. $79.84, *b* = $52.09, 95% CI [47.78, 56.40]; all *p*s < .001).

The difference in willingness to pay between the Human and AI therapists was significantly larger in the Fully Autonomous AI condition than in the Assistive AI condition at the trainee (*b* = -$9.47, 95% CI [-18.03, −0.92], *t* = −2.17, *p* = .030), master’s (*b* = -$15.68, 95% CI [-24.07, −7.29], *t* = −3.66, *p* < .001), and doctoral (*b* = -$20.09, 95% CI [-28.71, −11.47], *t* = −4.57, *p* < .001) levels, but not at the peer-support level. Similarly, the difference between Human and AI ratings was significantly larger in the Fully Autonomous than in the Collaborative AI condition at all experience levels: peer support (*b* = - $8.21, 95% CI [-14.30, −2.11], *z* = −2.64, *p* = .008), trainee (*b* = -$14.65, 95% CI [-20.65, −8.65], *z* = −4.79, *p* < .001), master’s (*b* = -$22.02, 95% CI [-27.78, −16.26], *z* = −7.50, *p* < .001), and doctoral (*b* = -$23.66, 95% CI [-29.75, −17.58], *z* = −7.62, *p* < .001). There was no significant differences between the Collaborative and Assistive AI conditions at any level of therapist experience (all *p*s > .05).

#### 3.2.3 Willingness to wait by therapist experience level

##### Refusal

Pairwise comparisons revealed that participants were significantly more likely to refuse the AI therapist than the Human therapist across all 12 combinations of therapist experience and AI integration (all *ps* < .001; see **Figure 3**). For the Human therapist, refusal rates were highest at the peer-support level (15.3%), compared to the trainee (3.3%), master’s (1.0%), or doctoral level (1.2%). Refusal rates for Assistive and Collaborative AI decreased ranged from 15% to 24%, with highest refusal rates for peer experience levels. Refusal rates for Fully Autonomous AI were relatively similar at different comparison therapist experience levels, ranging from 29% to 32%.

##### Wait duration

Among participants who indicated that they would be willing to see the therapist, wait duration was analyzed using a cumulative link mixed model. Pairwise comparisons revealed that participants were willing to wait significantly longer for the Human therapist than for the AI therapist across all 12 combinations of therapist experience and AI integration (all *ps* < .001). See **Figure 3**.

#### 3.2.4. Likelihood, hesitancy, and acceptability by therapist experience level

##### Likelihood

Participants rated the Human therapist as significantly more likely to be chosen than the AI therapist across all 12 combinations of AI integration and therapist experience (all *p*s < .001). The difference in likelihood ratings between Human and AI therapists was significantly larger in the Fully Autonomous AI condition than in the Assistive AI condition at every experience level: peer support (*b* = - 0.29, 95% CI [-0.49, −0.10], *t* = −2.94, *p* = .003), trainee (*b* = −0.36, 95% CI [-0.63, −0.08], *t* = −2.54, *p* = .011), master’s (*b* = −0.66, 95% CI [-0.93, −0.39], *t* = −4.77, *p* < .001), and doctoral (*b* = −0.66, 95% CI [- 0.94, −0.38], *t* = −4.68, *p* < .001) levels. Similarly, this difference was significantly larger in the Fully Autonomous than in the Collaborative AI condition at the trainee (*b* = −0.58, 95% CI [-0.77, −0.39], *z* = - 5.91, *p* < .001), master’s (*b* = −0.83, 95% CI [-1.02, −0.65], *z* = −8.83, *p* < .001), and doctoral (*b* = −0.79, 95% CI [-0.98, −0.59], *z* = −7.91, *p* < .001) levels, but not at the peer-support level (*b* = −0.16, 95% CI [- 0.36, 0.03], *z* = −1.64, *p* = .101). There were no significant differences between the Collaborative and Assistive AI conditions at any experience level (all *p*s > .05).

##### Hesitancy

Participants reported significantly greater hesitancy toward the AI therapist than the Human therapist across all 12 combinations of AI integration and therapist experience (all *p*s < .001). The difference in hesitancy ratings between Human and AI therapists was significantly more in the Fully Autonomous AI condition than in the Assistive AI condition at every experience level: peer support (*b* = 0.35, 95% CI [0.16, 0.54], *t* = 3.55, *p* < .001), trainee (*b* = 0.39, 95% CI [0.12, 0.65], *t* = 2.82, *p* = .005), master’s (*b* = 0.61, 95% CI [0.35, 0.88], *t* = 4.56, *p* < .001), and doctoral (*b* = 0.65, 95% CI [0.38, 0.92], *t* = 4.68, *p* < .001) levels. Similarly, this difference was significantly larger in the Fully Autonomous than in the Collaborative AI condition at all experience levels: peer support (*b* = 0.24, 95% CI [0.05, 0.43], *z* = 2.47, *p* = .014), trainee (*b* = 0.63, 95% CI [0.44, 0.82], *z* = 6.58, *p* < .001), master’s (*b* = 0.88, 95% CI [0.70, 1.06], *z* = 9.55, *p* < .001), and doctoral (*b* = 0.89, 95% CI [0.70, 1.09], *z* = 9.18, *p* < .001). There were no significant differences between the Collaborative and Assistive AI conditions at any experience level (all *p*s > .05).

##### Acceptability

Participants rated the Human therapist as significantly more acceptable than the AI therapist across all 12 combinations of AI integration and therapist experience (all *p*s < .001). The difference in acceptability ratings between Human and AI therapists was significantly larger in the Fully Autonomous AI condition than in the Assistive AI condition at every experience level: peer support (*b* = - 0.36, 95% CI [-0.54, −0.17], *t* = −3.81, *p* < .001), trainee (*b* = −0.32, 95% CI [-0.58, −0.06], *t* = −2.42, *p* = .016), master’s (*b* = −0.64, 95% CI [-0.89, −0.38], *t* = −4.91, *p* < .001), and doctoral (*b* = −0.53, 95% CI [- 0.79, −0.27], *t* = −4.00, *p* < .001) levels. Similarly, this difference was significantly larger in the Fully Autonomous than in the Collaborative AI condition at all experience levels: peer support (*b* = −0.27, 95% CI [-0.45, −0.09], *z* = −2.87, *p* = .004), trainee (*b* = −0.58, 95% CI [-0.76, −0.40], *z* = −6.27, *p* < .001), master’s (*b* = −0.89, 95% CI [-1.06, −0.71], *z* = −9.97, *p* < .001), and doctoral (*b* = −0.80, 95% CI [-0.99, - 0.62], *z* = −8.55, *p* < .001). There were no significant differences between the Collaborative and Assistive AI conditions at any experience level (all *p*s > .05).

### 3.3 Vignette depression severity

There was no significant effect of vignette severity on therapist preference. Participants strongly preferred the Human therapist over the AI therapist at every severity level (all *ps* < .001), and there was no significant difference in preference levels across severity conditions (see **Figure S2**). In addition, vignette severity did not significantly moderate willingness to pay, willingness to wait, likelihood of seeking treatment, hesitancy, or acceptability (see **Figure S2**).

### 3.4 Individual-difference moderators

To examine whether individual differences moderated the effect of degree of AI integration on therapist preference, we estimated four models corresponding to demographic characteristics, clinical characteristics, AI experience, and access to care. Each model included the main effects of the individual-difference variables and their interactions with degree of AI integration. Bonferroni adjustments were applied within each model set. Finally, we estimated a combined model that included all individual-difference moderators. Full model results are presented in **Table S15**.

In the demographic model, age and gender significantly predicted overall therapist preference, with men showing greater overall preference for the AI therapist. Age was the only demographic characteristic that significantly interacted with degree of AI integration, with older participants showing relatively more favorable evaluations of Fully Autonomous AI. In the AI-experience model, AI self-efficacy, AI usage frequency, and prior use of AI for mental health significantly predicted overall therapist preference, with higher AI self-efficacy and more frequent AI use associated with greater preference for the AI therapist. AI self-efficacy, AI literacy, and prior use of AI for mental health significantly interacted with degree of AI integration. Higher AI self-efficacy and AI literacy were associated with relatively more favorable evaluations of Fully Autonomous AI. No variables were significant in the clinical-characteristics or access-to-care models after correction for multiple comparisons. See **Table S15** for test statistics.

In the combined model, only AI self-efficacy and AI usage frequency remained significant predictors of overall therapist preference. AI self-efficacy and AI literacy also remained significant moderators of the effect of AI integration, such that participants with greater AI self-efficacy and AI literacy evaluated Fully Autonomous AI more favorably.

## 4. DISCUSSION

Artificial intelligence (AI) is increasingly being incorporated into mental health care, ranging from AI tools that support clinicians to chatbots that interact directly with patients. As health care systems and companies integrate AI into psychotherapy, relatively little research has examined the perspectives of key stakeholders in treatment. To our knowledge, the present study is the first to examine how individuals with self-reported depression evaluate psychotherapy delivered by a human therapist without the use of AI compared with psychotherapy involving AI tools and chatbots.

The present study shows that participants consistently valued Human therapists – or therapists who did not use AI – more highly than AI therapists. Participants preferred the Human therapist, reported a greater likelihood of seeking treatment from the Human therapist, and rated treatment from the Human therapist as more acceptable. They also reported less hesitancy about seeking treatment from the Human therapist and were less likely to refuse treatment. In addition, participants were willing to pay more and wait longer for treatment from the Human therapist. These preferences for Human over AI therapy were particularly pronounced at higher levels of therapist experience. However, we did not observe that vignette depression severity moderated participants’ evaluations.

### 4.1. Degree of AI integration

Participants evaluated Assistive and Collaborative AI similarly across most outcomes. However, Fully Autonomous AI was evaluated substantially less favorably. Participants showed a stronger preference for the Human therapist when the alternative was Fully Autonomous AI and reported lower likelihood of seeking treatment, greater hesitancy, and lower acceptability for Fully Autonomous AI. They were also willing to pay approximately 31% less for Assistive or Collaborative AI than for the Human therapist and 57% less for Fully Autonomous AI. In addition, they were less willing to wait for Fully Autonomous AI and more likely to refuse it.

These findings extend prior research on the acceptability of AI-based digital mental health interventions. Békés and Aafjes-van Doorn ^1^ found that teletherapy was viewed more positively than AI chatbots and AI avatar-based interventions. The present findings similarly indicate that people continue to place greater value on psychotherapy delivered without AI involvement. However, our findings further suggest that evaluations depend on the degree to which AI is integrated into psychotherapy. Although Collaborative AI was described as having a greater role in treatment than Assistive AI, participants did not evaluate Collaborative AI less favorably. Thus, participants might be accepting of varying degrees of AI involvement as long as a human therapist remains directly involved in treatment. Evaluations become substantially less favorable when AI operated autonomously and assumed responsibility for treatment decisions.

This pattern was particularly pronounced at higher levels of therapist experience. At the doctoral level, participants were willing to pay approximately 65% less for Fully Autonomous AI than for the Human therapist. In addition, 36.1% of participants indicated a complete preference for the Human therapist over Fully Autonomous AI, compared with 23.5% when the alternative used Assistive AI. These findings suggest that a substantial proportion of participants were strongly opposed to psychotherapy delivered autonomously by AI.

Prior research suggests that autonomous or largely autonomous AI interventions can produce clinical benefits. Li et al. ^12^ found that AI-based conversational agents were associated with reductions in depression and psychological distress, and Heinz et al. ^13^ similarly found that Therabot produced greater symptom reductions than a waitlist condition, was well utilized, and received favorable ratings from users. However, participants in the latter study volunteered for a trial of an AI chatbot and therefore likely represent a subset of the population that is particularly open to AI-based treatment.

Interestingly, we observed evidence of a contrast effect: participants sometimes evaluated the Human therapist more favorably when the alternative was Fully Autonomous AI than when the alternative used Assistive AI. Because the description of the Human therapist remained the same across these comparisons, the less favorable evaluation of Fully Autonomous AI might have increased the perceived value of the Human therapist by comparison.

A similar pattern has been observed in judgments of AI-generated art. Bellaiche, et al. ^21^ presented participants with AI-generated artwork that was randomly labeled as either Human-created or AI-created and found that artwork attributed to a human was evaluated more favorably, particularly in terms of profundity and monetary value. Bellaiche et al. suggested that people might value art not only as a perceptual product, but also as a medium for communicating human experience. A similar distinction might apply to psychotherapy: patients might value psychotherapy not only for the outcomes it provides, but also for the human relationship through which treatment is delivered. From this perspective, AI integration might reduce the perceived value of psychotherapy when it is seen as replacing a core element of treatment that patients regard as distinctly human.

### 4.2. Therapist experience

Participants placed greater value on Human therapists with higher levels of experience. Therapist experience was also associated with greater willingness to pay for Assistive and Collaborative AI. However, willingness to pay for Fully Autonomous AI did not differ across therapist experience levels. Thus, greater therapist experience increased the perceived value of Human therapists and therapists who integrated AI into treatment, but not the perceived value of Fully Autonomous AI.

Differences in evaluations of the Human and AI therapists became larger at higher levels of therapist experience. Participants were relatively more willing to consider AI when the Human alternative was a peer-support provider and less willing when the Human alternative was a master’s or doctoral-level therapist. This pattern was particularly pronounced for Fully Autonomous AI, whose perceived value did not increase across therapist experience levels. Thus, Fully Autonomous AI appeared most competitive with peer support and least competitive with master’s- and doctoral-level Human therapists.

### 4.3. Vignette depression severity

Surprisingly, vignette depression severity did not moderate participants’ evaluations of the Human and AI therapists. The pattern of preferences was similar whether participants imagined experiencing mild, moderate, severe, or severe depression with suicidal ideation. Participants’ own depression and anxiety symptom severity also did not moderate the effect of AI integration.

One possible explanation is that participants placed greater emphasis on the description of the therapist than on the severity of the hypothetical symptoms. Each vignette comprised information about depression severity, therapist experience, provider type, and degree of AI integration. Participants might not have fully incorporated all of this information into their judgments.

### 4.4. Individual differences

AI experience predicted attitudes towards AI therapists. In the model including all moderators, greater AI self-efficacy and more frequent AI use predicted greater overall preference for the AI therapist. AI self-efficacy and AI literacy also moderated the effect of AI integration, such that participants with greater confidence in using AI and greater knowledge of AI evaluated Fully Autonomous AI more favorably relative to Assistive AI.

These findings are consistent with those of Békés and Aafjes-van Doorn’s^1^, who found that general attitudes toward AI were strongly associated with acceptance of AI chatbots and AI avatars. They also found that older age was associated with greater acceptance of AI-based interventions. Similarly, in the present study, age moderated preferences in the demographic model, with older participants evaluating Fully Autonomous AI relatively more favorably. However, this effect did not remain significant in the combined model. Gender and prior use of AI for mental health also predicted preferences in the grouped models but were no longer significant in the combined model. Future research should examine whether age, gender, and prior use of AI for mental health remain associated with attitudes toward AI-integrated psychotherapy after accounting for AI-experience characteristics.

### 4.5. Limitations

The present study examined hypothetical preferences rather than actual treatment decisions. Participants were recruited through Prolific and might have had greater familiarity with technology and online research than the general population. Nevertheless, participants strongly favored Human therapists across outcomes.

Vignette depression severity was manipulated between participants, whereas AI integration and therapist experience were manipulated within participants. Therefore, the study might have had less power to detect smaller effects of vignette depression severity. In addition, the severity description might not have been sufficiently salient relative to the other vignette information presented in the vignettes.

### 4.6. Implications

The present findings suggest that prospective patients might be more accepting of AI when it is used by a Human therapist than when it replaces one. Participants evaluated Assistive and Collaborative AI similarly, despite the greater role of AI in the Collaborative condition. However, they consistently evaluated Fully Autonomous AI less favorably. Accordingly, AI features such as guided intake, session summaries, documentation support, journaling prompts, and between-session exercises might be more acceptable to patients than treatment delivered autonomously by AI.

Mental health providers should nevertheless be cautious when integrating AI into their psychotherapy. At the doctoral level, therapists who used AI were assigned a lower monetary value than Human therapists at the trainee level and only a slightly higher value than Human peer-support providers. Thus, although patients might accept AI when a Human therapist directs treatment, they might perceive its use as reducing the value of the clinician’s expertise.

Health systems should also be wary about assuming that lower costs and immediate availability will be sufficient to encourage patients to use Fully Autonomous AI. Participants indicated that they were willing to pay less and wait less for autonomous treatment, but the present study did not directly evaluate whether lower costs or shorter wait times would compel them to choose Fully Autonomous AI over a Human therapist. Similarly, participants reported a lower willingness to pay for a human therapist using Assistive or Collaborative AI in their psychotherapy, which poses an open question as to why participants are willing to pay less for AI-integrated psychotherapy than fully human therapy.

### 4.7. Future directions

The present study assessed self-report preferences in hypothetical treatment scenarios. Future research could examine the factors that influence actual behavioral decisions to enroll in or avoid AI-integrated psychotherapy. As health care companies evaluate AI chatbots and AI-integrated psychotherapy services, these evaluations should include patient-reported outcomes in addition to symptom-based outcomes, such as treatment satisfaction, the perceived importance of symptom change, adverse effects, and adherence to treatment activities such as homework ^22–25^.

Future studies should also examine the acceptability of AI for specific psychotherapy activities. For example, patients might be comfortable with AI being used for notetaking but less comfortable with AI developing treatment plans or making clinical decisions. In addition, future studies should examine the characteristics and concerns of the subset of participants who strongly reject AI-integrated psychotherapy.

## Data availability statement

The data that support the findings of this study are available from the corresponding author upon reasonable request.

## Ethics Declaration

This study was approved by the University of Idaho Institutional Review Board (Protocol 26-068) and was conducted in accordance with the Declaration of Helsinki. All participants provided informed consent prior to participation.

## Acknowledgments

None.

